# Stakeholder Perspectives on the Feasibility, Acceptability, and Implementation Strategies for Adapted Autism Spectrum Disorder Screening Tools in Community Settings in Kilimanjaro, Tanzania

**DOI:** 10.64898/2026.09.12.26362904

**Authors:** Florida J. Muro, Alex Gabagambi Alexander, Catherine Mbughuni, Theonest Ndyetabura, Aloyce George Mlyomi, Rosalia Njau, Innocent Peter, Thierry Kabwe, Elia Swai, Blandina Mmbaga, Jovin R Tibenderana

## Abstract

**Background:** Early identification of Autism Spectrum Disorder (ASD) is critical for optimizing developmental outcomes, yet community-based screening remains underutilized in Tanzania due to systemic, cultural, and logistical barriers.

**Purpose:** This study explores stakeholder perceptions of the acceptability, feasibility, and potential implementation strategies for culturally adapted autism screening tools in community-based early childhood settings in the Kilimanjaro region, Tanzania.

**Methods:** A phenomenological qualitative study guided by the Socioecological Model (SEM) was conducted across preschools, daycare centers, and rehabilitation facilities in Moshi Municipality and Hai District. Multi-stakeholder participants included daycare center directors, kindergarten teachers, healthcare providers, and parents and guardians of children with ASD and policymakers (10 in-depth interviews, 3 focus group discussions). Data were collected through usability assessments, cognitive debriefing, and semi-structured discussions, and were thematically analyzed using NVivo 15 aligned with SEM domains.

**Results:** Participants reported high perceived acceptability of utilizing the tools, driven by the tools’ Swahili adaptation, perceived clinical usefulness, and cultural relevance. Key anticipated feasibility enablers included private administration of the tools to mitigate stigma, trained non-specialist screeners, and structured caregiver psychoeducation. Primary barriers included questionnaire length, caregiver stress, and initial resistance. Multi-level implementation strategies emerged: standardized training, consistent home–school communication, multidisciplinary care coordination, task-shifting to Community Health Workers, engagement of local and faith leaders, and integration into digital and policy frameworks.

**Conclusion:** Culturally adapted ASD screening tools are feasible and acceptable in Tanzanian community settings when deployed with attention to linguistic accessibility, confidentiality, staff capacity, and cross-sectoral partnerships. Embedding screening within existing early childhood and community health infrastructure, supported by policy alignment and digital outreach, offers a scalable model for improving early ASD identification in low-resource contexts.

## Background

Early identification of Autism Spectrum Disorder (ASD) is essential, as timely intervention during sensitive developmental windows can significantly improve long-term communication, adaptive functioning, and quality of life (Okoye *et al*., 2023). ASD is a common neurodevelopmental condition and an increasing public health concern. It affects approximately 61.8 million individuals, or one in every 127 people worldwide(Santomauro *et al*., 2024; World Health Organization, 2025). While the estimated prevalence of ASD in Sub-Saharan Africa is approximately 1%, the true burden in Tanzania remains largely unquantified due to systemic under-identification and weak surveillance systems in the community (Abubakar *et al*., 2016; Kantawala *et al*., 2023). These disparities are particularly pronounced in peri-urban and low-income communities, where access to paediatric specialists is limited, cultural stigma surrounding developmental differences persists, and formal screening infrastructure is absent (Manji and Hogan, 2013). In this context, preschools, daycare centres, and family networks represent important entry points for early detection of ASD(Zwaigenbaum *et al*., 2019). These trends underscore a significant public health gap. Even when tools are culturally adapted, their practical uptake is frequently hindered by high staff-to-child ratios, limited administrative capacity, and caregiver time burdens.

Failure to ensure that screening tools are feasible and acceptable can undermine their uptake, effectiveness, and sustainability, ultimately limiting their public health impact. This is because tools perceived as overly lengthy, culturally misaligned, or administratively burdensome are unlikely to be adopted, thereby perpetuating diagnostic delays and missed intervention windows (Damschroder *et al*., 2022; Sekhon *et al*., 2017; Zwaigenbaum *et al*., 2019). As a result, children with possible ASD or other developmental concerns may remain unidentified, ultimately limiting timely access to early intervention services. Early intervention services and identification enable access to evidence-based early support, which is strongly associated with improved cognitive functioning, reduced behavioural challenges, and greater independence in adulthood (Gabbay-Dizdar *et al*., 2022). Whenever there are delays in identification of ASD, their impact extends beyond the child to the wider family and community.

In the absence of feasible and acceptable community screening mechanisms, children with ASD often experience prolonged educational exclusion, heightened caregiver stress, and increased lifelong dependency, placing substantial burden on families and national health and education systems(Okoye *et al*., 2023). These combined pressures not only undermine household wellbeing but also contribute to long-term dependency, thereby increasing the burden on health and education systems. Addressing the anticipated feasibility and perceived acceptability of tools is therefore essential. Feasible and acceptable screening may facilitate earlier identification and referral, as this is critical for supporting optimal developmental trajectories and fostering inclusive community development across Tanzania.

Facility-based studies have documented clinical characteristics, caregiver knowledge, and service gaps (Harrison *et al*., 2014). Moreover, community initiatives led by non-governmental organizations, such as those of Autism Beyond Borders Tanzania (ABBTZ) and Connects Autism Tanzania (CAT), have introduced awareness campaigns and educator training in urban centres. Building on these local efforts, there is also growing evidence supporting the use of standardized international screening tools in similar resource-limited contexts. Globally validated tools such as the Modified Checklist for Autism in Toddlers, Revised with Follow-Up (M-CHAT-R/F) and the Social Communication Questionnaire (SCQ) have demonstrated strong psychometric properties across diverse settings and have been adapted for use in several African countries, including Mali, Kenya, and South Africa(Kipkemoi *et al*., 2025; Sangare *et al*., 2019; Vorster *et al*., 2021). Importantly, these instruments serve different developmental age groups and screening purposes. The M-CHAT-R/F is primarily designed for toddlers aged approximately 16–30 months and focuses on identifying children who may be at elevated likelihood of ASD for further evaluation. In contrast, the SCQ is designed for older children, generally from 4 years of age, and assesses social communication and related behaviours associated with ASD. Their complementary age ranges make them potentially useful for extending community-based screening across different stages of early childhood. However, their successful implementation depends not only on psychometric performance but also on their feasibility, acceptability, and fit within routine community and early childhood settings. The M-CHAT-R/F (20 items; children aged 16–30 months) and the SCQ (40 items; children aged 4 years and older) were adapted for the Tanzanian context through forward and backward translation between English and Kiswahili, expert panel review, and cognitive debriefing with caregivers, following ISPOR good-practice principles (Wild *et al*., 2005)

Despite incremental progress, critical implementation gaps persist. There is a notable absence of robust, multi-stakeholder exploration of the real-world feasibility and perceived acceptability of culturally adapted ASD screening tools in Tanzanian early childhood settings. As a result, previous initiatives have not consistently examined anticipated feasibility barriers such as time burden, training requirements, privacy concerns, stigma-related reluctance, and linguistic comprehension across diverse ecological levels. Without rigorous anticipated feasibility and perceived acceptability data about the tools, early identification efforts remain fragmented, culturally misaligned, and unsustainable.

This study aims to explore stakeholders’ perceptions of feasibility, acceptability, and implementation strategies of culturally adapted ASD screening tools (M-CHAT-R/F and SCQ) guided by the Socioecological Model (SEM) in community-based early childhood settings in the Kilimanjaro region, Tanzania.

## Methods

### Study Design

A qualitative descriptive study design was used to explore stakeholders’ perceptions regarding the feasibility, acceptability, and implementation of culturally adapted ASD screening tools in community settings. The study was guided by the SEM, which provided a framework for examining factors influencing implementation at individual, interpersonal, community, and institutional levels.

### Study Setting and Population

The study was conducted in selected preschools, daycare centres, and community rehabilitation facilities across Moshi Municipality and Hai District in the Kilimanjaro Region, namely Gabriella Rehabilitation Centre, Faraja Primary, and Mwereni Primary School. The target population includes primary caregivers of children aged 16 months to 8 years, preschool teachers, daycare providers, centre administrators, community health workers (CHWs), local leaders, and district-level education/health officials.

### Sampling Strategy

Early childhood sites in Moshi and Hai districts were identified through snowball sampling to facilitate access to diverse urban and peri-urban settings. Within the identified sites and relevant community networks, participants were purposively recruited based on their roles and experiences relevant to the anticipated feasibility and perceived acceptability of ASD screening. Stakeholder groups included caregivers, educators, healthcare and allied health professionals, and other relevant community stakeholders. Recruitment continued iteratively until thematic sufficiency was achieved within and across stakeholder groups.

### Data Collection

We conducted 10 in-depth interviews (IDIs), selecting participants from each of the five key stakeholder categories: daycare center directors, kindergarten teachers, healthcare providers, and parents and guardians of children with ASD and policymakers. In addition, we conducted 3 FGDs with five participants from each group by using purposive sampling to gather group insights from the same stakeholder categories to capture shared experiences, attitudes, and contextual insights that may not emerge in one-on-one interviews. Both interview guides were piloted within 3 preschools. All IDIs and FGDs were guided by a semi-structured interview guide, allowing for consistency across interviews while also enabling flexibility to explore participant-specific experiences and perspectives. Data collection continued until no substantively new themes emerged across the major stakeholder categories, indicating thematic saturation. This was conducted in Kiswahili by a trained research assistant with experience in qualitative data collection. Participants were provided with information about the study before being asked for informed consent. Interviews and discussions were conducted in a private room to ensure confidentiality and privacy. Data collection using a semi-structured guide lasted for around 45-60 minutes. Recordings were transcribed verbatim before translation into English.

### Data analysis

Audio recordings were transcribed verbatim and translated by the author from Swahili to English. All transcripts were read in full to facilitate familiarization with the data before being analyzed using a hybrid thematic analysis approach that combined deductive and inductive coding which was done by using NVivo version 15. A collaborative coding strategy using a codebook was developed by two independent coders with backgrounds in early childhood development conditions to help reduce the possibility of bias and ensure that the participants’ voices were accurately reflected in the analysis. An initial coding framework was developed deductively from the study objectives, interview guides, and the Socioecological Model, while additional codes were generated inductively from participants’ accounts. The researchers initially independently coded a sample of interview transcripts, then compared their interpretations and refined the codebook through discussion. Differences in coding were resolved by consensus, and where necessary, in consultation with a senior researcher experienced in qualitative research. An audit trail documenting coding decisions, codebook revisions, memos, and theme development was maintained throughout the data analysis, while reflexive notes were used to consider how researchers’ professional backgrounds and assumptions might have influenced interpretation. The authors reviewed and named the themes clearly as per SEM. Finally, produce a report that presents a coherent narrative of the themes based on the study objectives.

### Ethical Considerations

Ethical approvals were obtained from the KCMC University Research Ethics Review Committee (KURERC) with certificate number 2789 and the National Health Research Ethics Committee (NatHREC). Written informed consent was obtained from all adult participants. To mitigate stigma and emotional distress, screening is conducted in private settings, data are anonymized using unique identifiers, and participants may withdraw at any time. All digital data was encrypted and stored on password-protected servers in compliance with Tanzanian data protection guidelines. Participants received modest compensation for transport and time in accordance with ethical standards.

## RESULTS

The study participants were predominantly female (90%) with a median age of 33 years (IQR: 28–39). Most participants had attained at least a diploma or bachelor’s level education, and represented a range of roles including health and allied professionals, special education teachers, caregivers, NGO staff, and institutional and rehabilitation center leaders. A significant proportion of participants (80%) reported prior training in both child development and autism or developmental delay (Table 1). Table 2 describes the characteristics of study participants in Focus Group Discussions.

**Table 1.** Sociodemographic Characteristics of IDI Study Participants (N = 10)

| Characteristic | Category | Frequency<br>/median | Percent/IQR |
| --- | --- | --- | --- |
| **Median Age (years)** | Median (IQR) | 33 | (28–39) |
| **Sex** | Female | 9 | 90.0 |
|  | Male | 1 | 10.0 |
| **Education Level** | Primary Education | 2 | 20.0 |
|  | Diploma | 3 | 30.0 |
|  | Bachelor Degree | 4 | 40.0 |
|  | Master Degree | 1 | 10.0 |
| **Occupation** | Health/Allied Professionals* | 2 | 20.0 |
|  | Special Education Teachers | 2 | 20.0 |
|  | NGO Representative | 1 | 10.0 |
|  | Policy Maker | 1 | 10.0 |
|  | Directors | 2 | 20.0 |
|  | Caregivers/Parents | 2 | 20.0 |
| **Residence/Workplace** | Urban | 7 | 70.0 |
|  | Rural | 3 | 30.0 |
| **Child Development Training** | Yes | 8 | 80.0 |
|  | No | 2 | 20.0 |
| **Autism/Developmental Training** | Yes | 8 | 80.0 |
|  | No | 2 | 20.0 |
| **Heard of Autism Spectrum** | Yes | 9 | 90.0 |
|  | No | 1 | 10.0 |
| **First Source of Autism Information** | School | 3 | 30.0 |
|  | Health Facility | 4 | 40.0 |
|  | Other Sources | 3 | 30.0 |

**Table 2.** Sociodemographic Characteristics of Study Participants in FGDs (N = 15)

| **Characteristic** | **FGD 1: Occupational Therapists (n=5)** | **FGD 2: Parents/Caregivers (n=5)** | **FGD 3: Teachers (n=5)** |
| --- | --- | --- | --- |
| **Age (years)** | Median (IQR): 32 (28–44) | Median (IQR): 40 (27–60) | Median (IQR): 47 (38–50) |
| **Sex** | Female: 3 (60%) Male: 2 (40%) | Female: 4 (80%) Male: 1 (20%) | Female: 5 (100%) |
| **Education Level** | Diploma: 3 (60%)<br>Bachelor: 2 (40%) | Primary/Lower: 3 (60%)<br>Diploma: 1 (20%)<br>Bachelor 1 (20%) | Diploma: 4 (80%)<br>Bachelor: 1 (20%) |
| **Occupation/Role** | Occupational Therapist (100%) | Parent/Caregiver (100%) | Teacher (100%) |
| **Work Setting** | Private: 2 (40%)<br>Government: 2 (40%)<br>FBO: 1 (20%) | Informal/household-based | School-based (100%) |
| **Years of Experience** | Median (IQR): 6 (3–13) | N/A (caregiving experience varies) | Median (IQR): 4 (3–5) |
| **Child Development Training** | Yes: 100% | Yes: 40% No: 60% | Yes: 40% No: 60% |
| **Autism Training** | Yes: 100% | Yes: 30% No: 50% Not sure: 20% | Yes: 20% No: 80% |
| **Heard of Autism Spectrum** | Yes: 100% | Yes: 80% No: 20% | Yes: 100% |
| **First Source of Autism Information** | School/Radio | School/Health Facility/Other | Health Facility |

### Perceived acceptability and Anticipated feasibility of Using the Tools

#### Individual Level: Perceptions and Psychological Readiness

The Positive Perception of Tool Usefulness was reported to be a significant driver of individual perceived acceptability. Stakeholders, particularly educators, perceive these screening tools as essential instruments for identifying a child’s specific functional limitations and strengths. Participants demonstrated high psychological readiness to adopt the Swahili-adapted M-CHAT-R/F and SCQ tools.

> “I think it would be a good tool because it would help me understand the child’s challenges…” (FGD3, Teachers).

Furthermore, perceived acceptability often increases through personal engagement with the tool; while parents may initially show resistance, they frequently transition toward acceptance as they recognize their child’s behaviors within the questionnaire items. According to one specialist,

> “Sometimes a person may initially be resistant, but once they read the questions, they may begin to relate them to their own child… So automatically, they will start to accept it” (IDI1 special needs teacher).

However, the time burden in administration remains a challenge, as the psychological state and high stress levels of parents can hinder their concentration during long screenings. A special education expert highlighted that;

> “One challenge could occur when screening parents who have children with autism. Their patience may sometimes be limited… due to the child’s behaviour [or] the stress they are experiencing” (IDI5 Special Education).

#### Interpersonal Level: Communication and Confidentiality

The perceived acceptability and anticipated feasibility of the ASD screening process at the interpersonal level were found to be significantly shaped by the quality of engagement between the screener and the caregiver. Within the SEM, the interpersonal domain encompasses the dyadic interactions between caregivers and screening personnel relationships that directly influence trust, disclosure, and willingness to participate. Stakeholders emphasized that the manner in which screening questions were introduced and discussed could either facilitate or hinder parental engagement. When screeners adopted a warm, empathetic, and collaborative approach, caregivers reported feeling respected and psychologically safe, which in turn encouraged more honest and comprehensive reporting of their child’s developmental behaviors. Conversely, a rigid or clinical tone was associated with heightened anxiety, defensiveness, or reluctance to disclose concerns, particularly in contexts where developmental differences may be stigmatized or misunderstood. As one stakeholder articulated during an In-Depth Interview:

> “The manner in which someone engages a parent is important; if the approach is friendly and supportive, parents are more likely to respond openly” (IDI6, NGO representative).

Beyond communication style, the physical and social context in which screening questions are administered emerged as a critical determinant of perceived acceptability at the interpersonal level. Participants consistently emphasized that privacy during questioning was essential to maintain confidentiality, minimize social discomfort, and encourage open responses, particularly when discussing sensitive topics related to child development and potential neurodevelopmental concerns.

> “Sometimes parents find it difficult to say openly that their child has a problem. It feels like revealing something they do not want to announce publicly.” (FGD2, Caregivers)

In the Kilimanjaro context, where concerns about stigma surrounding developmental differences remain salient, caregivers expressed heightened sensitivity to being observed or overheard during screening. The fear of unintended disclosure, whether to neighbors, other parents, or community members, was identified as a potential barrier to full engagement with the screening process. As one caregiver articulated during a Focus Group Discussion:

> “It would be easier if the questions were asked privately between two people. It would be uncomfortable if the questions were asked in front of many people” (FGD2, Caregivers).

#### Organizational Level: Training and Administrative Anticipated feasibility

The importance of trained administrators emerged as a dominant factor influencing the anticipated feasibility of the screening program. Stakeholders, including preschool directors and caregivers, argued that for the culturally adapted M-CHAT-R/F and SCQ tools to be effective, those administering them must possess a deep understanding of the content and the underlying developmental concepts before engaging families.

Participants highlighted that without adequate training, there is a risk of misinterpretation of screening items, which could lead to either unnecessary parental anxiety or missed referrals. The organizational capacity to facilitate these trainings through workshops, ongoing supervision, and resource allocation was viewed as a critical precondition for successful implementation. Stakeholders noted that training not only enhances technical competence but also legitimizes the screening process in the eyes of parents, fostering trust in the institution administering the tool. As one caregiver remarked regarding the need for competent administration

> “First, the people administering the tools should be well trained so they understand them before explaining them to others” (FGD2, Caregivers).

Participants noted that when caregivers or teachers were initially informed about the number of items in the adapted M-CHAT-R/F or SCQ tools, concerns about completion time frequently arose. This apprehension was not merely about the minutes required to answer questions, but also about the cognitive and emotional energy needed to engage thoughtfully with developmental screening content amid competing priorities

> “The main challenge is time… when you tell them there are so many questions, they get discouraged and start wondering, how long will it take to finish?” (FGD1, OT).

#### Community and Policy Level: Linguistic and Cultural Adaptation

The availability of the adapted M-CHAT-R/F and SCQ tools in Swahili emerged as a critical factor for broad comprehension and uptake. Stakeholders emphasized that linguistic accessibility reduces barriers for non-specialist administrators, such as preschool teachers and daycare providers, and ensures that caregivers can engage meaningfully with screening content

> “Another thing I appreciate is that they are written in Swahili and are understandable. If they were in English, it would be a bigger problem” (FGD1, OT).

From a cultural adaptation standpoint, stakeholders highlighted that the absence of items addressing private or stigmatized behaviors, such as certain sensory-related questions, enhanced the tools’ social perceived acceptability. In contexts where discussions of bodily behaviors may be considered sensitive, this deliberate omission helped mitigate potential discomfort or resistance. As one preschool director observed;

> “Questions related to sensory behaviors like playing with private parts… are not included. So I do not think the questions will create serious cultural sensitivity issues” (IDI3, DR).

### Strategies for Improving Early Identification of ASD

#### Individual and Interpersonal Level: Empowerment and Consistency

Findings highlight that community education initiatives play a transformative role in shifting caregivers’ experiences from initial confusion or hopelessness toward informed advocacy. In the Kilimanjaro context, where awareness of ASD remains limited, and stigma may delay help-seeking, structured psychoeducation is reported to helps caregivers reframe their understanding of their child’s behavior and developmental needs. This process builds parental self-efficacy, enabling them to engage more confidently with screening and referral pathways.

> “…….. after spending time with them and providing education and counseling… they sometimes come back and say, ‘I’m sorry, I didn’t understand before. Now I understand my child.’ Once they reach that point, they often start educating others” (IDI1, Special needs teacher).

At the interpersonal level, stakeholders emphasized the importance of consistent communication and coordinated approaches across all adults involved in a child’s care, including parents, preschool teachers, daycare providers, and extended family members. Participants noted that using the same approach across caregivers helps create consistency in the child’s learning and daily interactions.

> “Everyone involved in educating the child must use the same approach or ‘language’” (IDI1, SNT).

#### Organizational Level: Multidisciplinary and Integrated Care

The Multidisciplinary Team (MDT) approach emerged as a key strategy for ensuring comprehensive assessment and coordinated support for children at risk of ASD. By bringing together professionals from diverse fields such as occupational therapy, education, and pediatrics. Organizations in Kilimanjaro can integrate multiple perspectives into a unified care plan. This collaborative model strengthens referral pathways and reduces fragmented service delivery, which is critical in resource-limited settings. As one teacher described:

> “All departments sit together and discuss: the occupational therapist has observed this, the teacher has observed that, the doctor has identified this… After that, we sit together and develop a plan” (IDI1, SNT).

To address constraints in specialized equipment, organizations reported actively improvising therapeutic tools using locally sourced materials. They suggested that this adaptive strategy enhances anticipated feasibility and sustainability, allowing educators and occupational therapists to deliver supportive interventions even without imported resources. For example, weighted jackets or sensory items crafted from local fabrics were cited as practical alternatives.

> “Children with autism require specialized materials, so we often improvise and create some tools using locally available materials” (IDI4, Director 4).

#### Community Level: Advocacy and Local Leadership

Some participants also reported that engaging local leadership emerged as a vital strategy for extending the reach of ASD screening to families in remote or underserved areas of Moshi and Hai districts. Stakeholders emphasized that village chairpersons, village executive officers, and ward executive officers possess intimate knowledge of household dynamics and can serve as trusted intermediaries between health/education systems and families. Building the capacity of these leaders to recognize early developmental concerns and to approach families with sensitivity was viewed as a sustainable pathway to improving case identification and reducing stigma.

> “One method we currently use is to work with village chairpersons, village executive officers, and ward executive officers. After we train them and build their capacity, they can help identify families in their communities who may have children with such challenges. They often know the families and can say, (IDI1, SNT).

Furthermore, some respondents reported that faith-based institutions emerged as a powerful platform for promoting early ASD identification and reducing stigma in Moshi and Hai districts. In Tanzania, where religious leaders hold significant moral authority and community trust, participants identified pastors, imams, and other faith leaders as influential ambassadors capable of normalizing developmental discussions within congregations. They suggested that by integrating ASD awareness into sermons, prayer groups, and community outreach, religious leaders can help reframe neurodevelopmental differences in culturally resonant, non-stigmatizing ways.

> “If we organize meetings with pastors and educate them… they can become excellent ambassadors. Through their sermons and prayers, they can help spread awareness” (IDI1, SNT).

Additionally, the participants revealed that integration of CHWs into ASD screening efforts emerged as a critical strategy for extending reach to underserved and remote populations in Moshi and Hai districts. CHWs, who routinely engage households through maternal-child health programs, immunization campaigns, and developmental monitoring, are uniquely positioned to incorporate early ASD identification into existing community health workflows

> “Community health workers interact with many people and can reach larger populations, even more than some institutions” (IDI2, Physiotherapist).

#### Policy Level: Digital Outreach and Systemic Integration

Stakeholders identified digital dissemination as a strategic lever for scaling ASD awareness and early identification efforts across Tanzania. Given the growing public reliance on social media and mobile technologies, participants emphasized that integrating autism education into widely used digital platforms could significantly extend reach, particularly among younger caregivers and educators in Moshi and Hai districts.

> “Many people now rely heavily on social media… Increasing the amount of information shared through these platforms could help raise awareness significantly” (IDI2, Physiotherapist).

Beyond digital outreach, participants highlighted the importance of embedding ASD screening within existing government structures such as public preschools, district health offices, and national awareness events to ensure sustainability and policy alignment. This multi-channel strategy (digital + institutional) supports the study’s goal of creating a scalable, community-led framework that leverages Tanzania’s evolving health and education infrastructure.

> “We can also use national awareness days that are already scheduled for different conditions. For example, the upcoming Down Syndrome awareness day could be used as an opportunity…” (IDI1_special need)

## DISCUSSION

This study explored the perceived acceptability, anticipated feasibility, and implementation strategies for culturally adapted ASD screening tools (M-CHAT-R/F and SCQ) in community settings in Moshi and Hai districts, Kilimanjaro Region, Tanzania. Guided by the Socioecological Model (SEM), findings indicated perceptions of the tools, while also identifying important anticipated barriers related to training, time, privacy, stigma, caregiver engagement, and integration into existing community and early childhood services. Stakeholders proposed implementation strategies centered on capacity building, community sensitization, stakeholder engagement, and integration of screening into existing service delivery platforms.

Stakeholders reported that the perceived usefulness of the adapted tools was a significant driver of perceived acceptability at the individual level. Educators and caregivers viewed the Swahili-adapted M-CHAT-R/F and SCQ as practical instruments for identifying children’s functional strengths and limitations, aligning with evidence that parent and teacher reported screeners are acceptable when they resonate with users’ lived experiences(Huda *et al*., 2024). Importantly, caregivers shifted toward acceptance as they recognized their child’s behaviors reflected in questionnaire items, a pattern consistent with research showing that engagement with screening content can foster insight and reduce denial (Franz *et al*., 2017).

At the interpersonal level, a supportive, non-judgmental communication style emerged as critical for encouraging honest disclosure. This finding reinforces the SEM premise that dyadic interactions directly shape health behaviors (Kenneth R Mcleroy, *et al*., 1988). Moreover, the strong preference for private administration underscores the salience of stigma in Tanzanian communities, where concerns about unintended disclosure may deter help-seeking (Kantawala *et al*., 2023). These results align with broader literature indicating that confidentiality safeguards and stigma-sensitive communication are essential for the perceived acceptability of developmental screening in low-resource settings (Faruk *et al*., 2020).

The necessity of trained administrators was a dominant organizational theme. Stakeholders emphasized that technical competence and conceptual understanding among screeners legitimize the process and build parental trust. This finding is supported by studies showing that training non-specialists improves screening fidelity and referral accuracy in LMICs(Naithani *et al*., 2022). This finding suggests that a screener’s role extends far beyond mere data collection, they must act as empathetic guides capable of navigating sensitive developmental discussions without triggering caregiver defensiveness or anxiety. Without adequate conceptual training, there is a tangible risk of misinterpreting screening items, which could lead to missed referrals or unnecessary psychological distress for families. The practical implication is that implementation efforts must prioritize standardized, Swahili-language training programs for non-specialist screeners (such as preschool teachers and community health workers) that integrate technical scoring skills with empathetic communication, stigma reduction, and clear referral guidance. From a policy perspective, this underscores the need for district health and education authorities to allocate dedicated resources and embed ASD-specific capacity building into existing professional development frameworks to ensure sustainable, high-quality screening.

At the same time, concerns about questionnaire length and time burden highlight a persistent tension between methodological rigor and practical anticipated feasibility. These challenges may be particularly pronounced in resource-constrained preschools where staff-to-child ratios are high. In such contexts, brief and modular tools with clear administration protocols are therefore recommended (Marguerite Marlow, *et al*., 2019).

The Multidisciplinary Team (MDT) approach was identified as a key strategy for comprehensive assessment. Integrating perspectives from occupational therapy, education, and pediatrics strengthens referral pathways and reduces fragmented care, a model increasingly advocated for ASD services in sub-Saharan Africa, contrasting with the MDT model’s goal of cohesive care, literature highlights that severe workforce shortages and disconnected medical-educational jurisdictions frequently perpetuate the very fragmentation these teams seek to eliminate, leaving a significant proportion of children without access to essential specialized therapies(Brewer, 2018). Consequently, while MDTs remain the ultimate clinical target, current evidence suggests that pragmatic transitional strategies, particularly task-shifting to community health workers and primary care providers, are critical to bridging this gap and ensuring immediate support in resource-constrained settings until a robust, specialized multidisciplinary workforce can be sustainably established(Franz *et al*., 2017)

Additionally, the creative improvisation of therapeutic materials using locally available resources demonstrates adaptive problem-solving that enhances sustainability in low-resource contexts(Aderinto *et al*., 2023).

The availability of tools in Swahili was universally valued as a prerequisite for comprehension and uptake. This supports international guidelines emphasizing that linguistic validation is not merely translation but requires cultural conceptual equivalence (Wild *et al*., 2005). Furthermore, the deliberate omission of culturally sensitive items (e.g., questions about private sensory behaviors) enhanced social perceived acceptability, illustrating the importance of participatory adaptation to align tools with local norms (Soto *et al*., 2015).

Community-level strategies, engaging village leaders, faith-based institutions, and Community Health Workers (CHWs), emerged as powerful levers for extending reach and reducing stigma. These findings confirm evidence that task-shifting to CHWs and leveraging trusted local influencers can effectively bridge gaps between formal health systems and underserved families (Feinberg and Eilenberg, 2024). At the policy level, digital dissemination via social media and integration into government structures were viewed as scalable mechanisms for awareness and systemic adoption. This multi-channel approach aligns with calls to embed developmental screening within existing maternal-child health and early education platforms to ensure sustainability (Coker *et al*., 2024).

The findings highlight important implications for practice, policy and research. In practice, implementation should prioritize standardized Swahili-language training for non-specialist screeners, integrating technical skills, communication, stigma reduction, and referral guidance (Naithani *et al*., 2022). Tool design should consider shorter or modular versions of adapted M-CHAT-R/F and SCQ to reduce time burden while maintaining validity, and screening must ensure confidentiality through private settings and clear data protection explanations (Marguerite Marlow, *et al*., 2019). From a policy perspective, community-based ASD screening can be integrated into Tanzania’s Early Childhood Development strategies and district health plans, leveraging existing community health worker and preschool systems (Coker *et al*., 2024). Policymakers should also promote digital health approaches, including mobile-based awareness in Swahili and allocate sustainable resources for training, supervision and referral systems through multi-sectoral collaboration (Martino and Naqvi, 2023). For research, future studies should focus on large-scale psychometric validation of adapted tools, assess long-term impacts on early diagnosis and child outcomes and conduct comparative studies across East Africa to strengthen culturally appropriate adaptation methods.

### Strengths and limitation

The study had several strengths and limitations. A key strength was its multi-stakeholder design guided by the Socio-Ecological Model, which enabled perspectives to be captured across different stakeholder levels and enhanced the contextual relevance of the findings. The study also examined both perceived acceptability and anticipated feasibility of culturally adapted ASD screening tools, providing contextually relevant insights into potential implementation considerations before wider community deployment.

However, data collection was confined to Moshi and Hai districts, which may limit the transferability of findings to other rural or regional contexts in Tanzania. In addition, feasibility was assessed based on stakeholders’ perceptions and anticipated implementation experiences rather than direct observation or measurement of screening activities under routine service conditions. Therefore, the findings may not fully reflect the actual time requirements, workflow demands, or operational challenges that could emerge during implementation. Finally, the study did not evaluate the diagnostic accuracy of the adapted tools against gold-standard clinical assessments; future research should assess their diagnostic performance and evaluate their feasibility and acceptability during real-world implementation.

## Conclusion

This study demonstrates that Stakeholders perceived culturally adapted M-CHAT-R/F and SCQ tools as potentially acceptable and feasible for use in selected community and early-childhood settings in Kilimanjaro when implemented with attention to multi-level factors outlined by the SEM. The important conditions for implementation include linguistic accessibility, stigma-sensitive communication, trained non-specialist administrators, strategic partnerships with stakeholders, and integration into existing health and education systems. However, these findings reflect stakeholder perceptions and should be confirmed through prospective pilot implementation assessing administration time, fidelity, caregiver uptake, screening outcomes, referral completion, and cost. Future efforts should prioritize large-scale validation and policy integration to sustain and expand these promising community-based approaches.

## Data Availability

All data produced in the present study are available upon reasonable request to the authors

## ACKNOWLEDGEMENT

The authors thankful for the support of Education Sub-Saharan Africa (ESSA) and the Research for Equitable Access and Learning Centre (REAL Centre) through Grant Reference ECDAFRICA001. This project was funded by ESSA and the REAL Centre with financial support from the Conrad N. Hilton Foundation and the Global Partnership for Education Knowledge and Innovation Exchange (GPE KIX). The authors also sincerely thank all study participants, experts, and stakeholders for their valuable time, contributions, and insights throughout the pilot implementation and adaptation process.

